# Not all animals are equal - farm living and allergy in Upper Bavaria

**DOI:** 10.1101/19007864

**Authors:** Matthias Wjst

## Abstract

**Background:** A lower allergy and asthma prevalence in farm children has been described three decades ago in Switzerland.

**Objective:** After years of research into bacterial exposure at farms, the origin of the farm effect is still unknown. We now hypothesize, that there is no such an effect in large industrial cattle farms with slatted floors indoors but in small farms only where animals are grazing outdoors and are having a higher endoparasite load.

**Methods:** We re-analyze an earlier epidemiological study by record-linkage to later agricultural surveys. The Asthma and Allergy Study in 1989/90 was a cross-sectional study of 1714 ten year old children in 63 villages covering ten different districts of Upper Bavaria. The farm effect is defined here as the association of number of cows per villager on lifetime prevalence of allergic rhinitis prevalence in the children of this village.

**Results:** The farm effect is restricted to small villages only. Furthermore, districts with higher Fasciola infection rates of cows, show a significant stronger farm effect than districts with lower infection rates.

**Conclusions:** The results warrant further research into human immune response to endoparasites in livestock.

## Introduction

The lower allergy prevalence in the farming population has been described already in one of the very first allergy text books in 1873 (1): „*These statistics of the occupations of hay-fever patients bring out prominently the very curious circumstance that the persons who are most subjected to the action of pollen belong to a class which furnished the fewest cases of the disorder, namely, the farming class*”. A direct allergy preventive effect by farming conditions remained unlikely as occupational asthma is frequently increased in farmers due to exposure to grain dust, animal dander and various chemicals (2).

In 1989, however, the „farm effect” was re-discovered by Gassner in the Canton of St. Gallen in the Swiss Alps (3), followed by studies of Braun-Fahrländer (4) and several other groups (5). We already noticed the effect 1989 in the Asthma and Allergy Study in Upper Bavaria, Germany (6) but attributed it to rather trivial reasons.

A healthy worker effect is likely in farmers, as allergy to grass or dust would leave to a drop-out of farming. Not unexpected farm children have a much lower family history of „hay fever” (4) possibly related to genetic differences in the farming population (7). Farm children may have experienced some kind of auto-desensitization due to the extreme allergen exposure (8). Furthermore, farmer’s children are known to supplement less vitamin D which means avoiding a known risk factor (9).

None of these objections have not been thoroughly addressed so far. While the „farm effect” has had little value in explaining the temporal rise of allergy, it may represent, however, an interesting observation where indeed an environmental substance may modify the allergic reaction. It has been reported several times that dust collected from cow stables has suppressive effects on the allergic sensitization in laboratory animals (10), (11). Although this could be an artifact as well either by the high allergen doses used or by the different immune reaction of mice, it raises the possibility that there is an indeed a protective factor of stable dust.

As IgE production in the human host is usually related to helminth exposure and not so much of bacterial load as being used as explanation (11 - 13), we now ask if cattle endoparasites could be responsible for an allergy modifying effect.

## Participants, materials and methods

Data of the Asthma and Allergy Study in Upper Bavaria 1989/1990 are used for an ecological re-analysis. Methods have been reported elsewhere including informed consent and ethics procedures (14 - 15). Briefly, we approached primary grade schools of 65 villages where 63 schools could visited in a semi-random order. 1714 of 1958 children participated in the study (87.5%). Parents filled in a self-administered questionnaire, children were tested by several allergens on the skin along with baseline lung function and lung function after inhalation of cold air. For the current analysis we did not any exclude any children.

Geocodes data for school districts have been obtained from geoportal.bayern.de/bayernatlas. Number of village inhabitants and assignment to districts was obtained of de.wikipedia.org/wiki/Oberbayern. The number of farm animals per village was imported from the official survey 1999 („Agrarstrukturerhebung”, www.statistikdaten.bayern.de/genesis, tables 41121-xxxx) where all farms with an area >2 ha or >8 cows are being recorded. Cow Fasciola infection ratio by districts was taken of a German thesis who tested between 2003 and 2005 milk samples from 80 farms in Bavaria for being positive in an anti-f2 Fasciola hepatica IgG assay. Altogether 5278 tank milk samples were examined and mean values calculated per district (16). Although most data here were acquired 10-15 years after the initial survey, farm size usually does not change and also conditions of wet farm ground with continuous re-infection remain unchanged. Life expectancy of cattle is around 20 years.

Data were visualized using the R libraries ggplot2, cross-tabulated and analyzed by logistic regression (17, 18).

## Results

Mean age of the children was 10 years with an age range of 9-13 years. There were 836 males, 870 females, while in 8 children sex was not known. Lifetime prevalence of physician diagnosed allergic rhinitis was 8.3% and of asthma 2.6%.

Supplemental Figure 1 shows the geographical distribution of the villages. As school sizes ranged from 6 to 63 children, mean rhinitis prevalence by village may be misleading in particular when villages are small.

Village size ranged from 858 to 8,578 residents, with a mean of 3,445 residents. Cow numbers ranged from 147 to 7,078 with a mean of 2,707 animals. The cows/resident ratio in the villages was found between 0 to 2.4 mit a mean of 0.9.

Allergic rhinitis prevalence decreased with increasing number of cows per village inhabitants, however, this effect was restricted to small villages (Supplement Figure 2) leading to the known 30%-40% reduction of allergic rhinitis.

Fasciola infection rate was found to be highly variable in the 10 districts with a prevalence between 18% and 98%. The prevalence of allergic rhinitis decreased with increasing Fasciola infection, however, only in those villages that have >1.5 cows/resident (Supplement Figure 3).

In a last step we analyze the data using separate logistic regression equations for each district (table). Fasciola infection was not significant on the p<0.05 level in any district while in the total sample the cows/resident ratio was a significantly protective factor (p=0.004, Figure).

**Figure:**
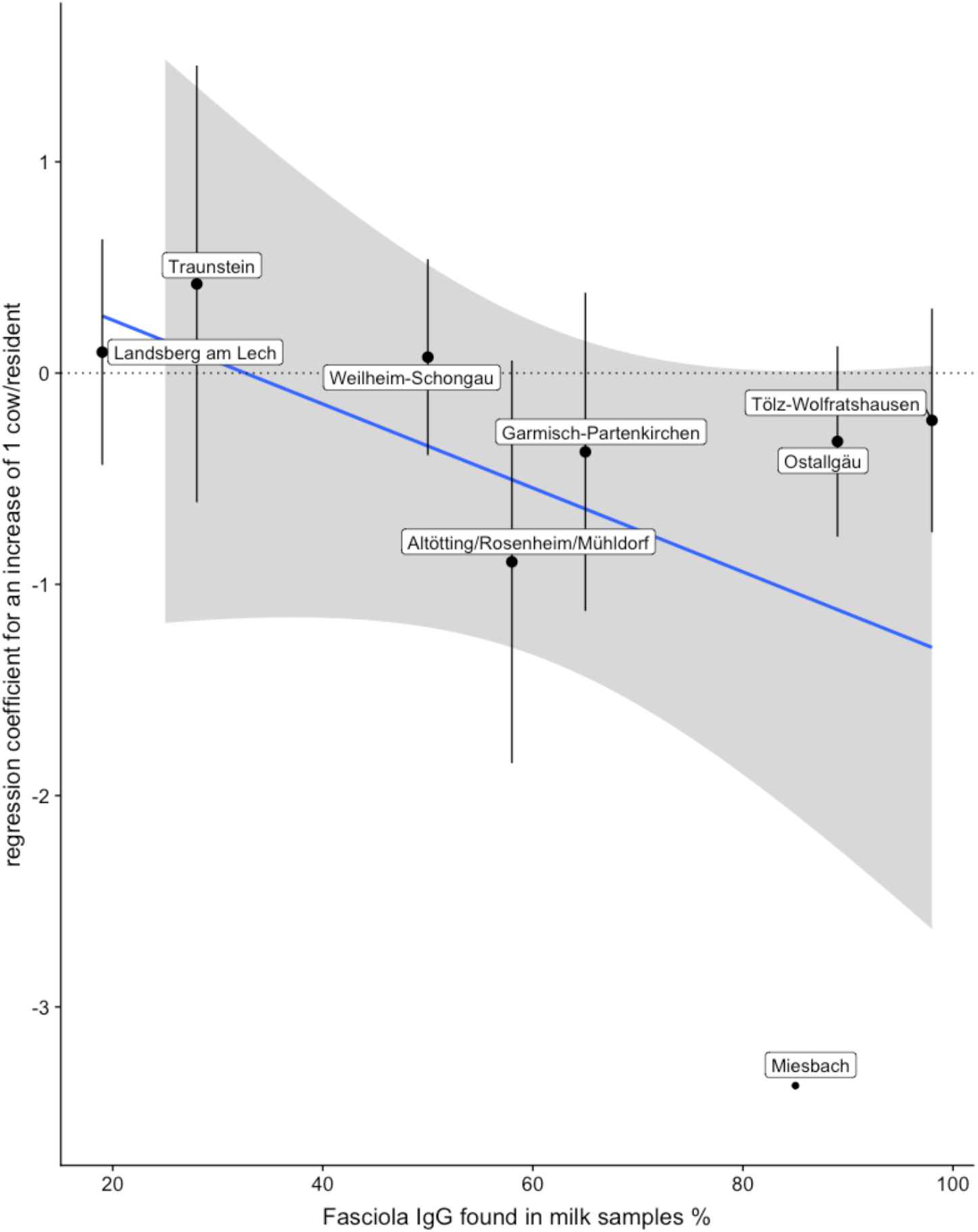
Logistic regression analysis of allergic rhinitis in Upper Bavaria 1989 in 1714 children. Regression coefficients from separate models for each district for the cows/resident ratio are plotted against the average percentage of milk samples with Fasciola IgG.

**Table:** Logistic regression analysis of allergic rhinitis in Upper Bavaria 1989 in 1714 children. Separate models are given for each district using the ratio of cows/resident as dependent variable. The three districts with the lowest number of children examined (Altötting N=19, Mühldorf N=77 and Rosenheim N=26 children) were collapsed into one category.

| district | % milk<br>samples<br>Fasciola<br>positive | children<br>N | regression<br>coefficient<br>cows/<br>resident | standard<br>error<br>cows/<br>resident | P |
| --- | --- | --- | --- | --- | --- |
| Altötting/Rosenheim/Mühldorf | 58 | 122 | -0,89 | 0,95 | 0,348 |
| Garmisch-Partenkirchen | 65 | 201 | -0,37 | 0,75 | 0,621 |
| Landsberg am Lech | 19 | 311 | 0,10 | 0,53 | 0,853 |
| Miesbach | 85 | 136 | -3,37 | 1,79 | 0,059 |
| Ostallgäu | 89 | 358 | -0,32 | 0,45 | 0,473 |
| Tölz-Wolfratshausen | 98 | 269 | -0,22 | 0,53 | 0,672 |
| Traunstein | 28 | 91 | 0,42 | 1,03 | 0,683 |
| Weilheim-Schongau | 50 | 226 | 0,08 | 0,46 | 0,871 |
| total sample | 64 | 1714 | -0,46 | 0,16 | 0,004 |

## Discussion

In this ecological analysis we replicate the known negative association of cattle farming and allergic rhinitis and find some preliminary evidence that cow endoparasites may be involved in this association. As there were no effects by other farm animals like pigs, goats, horses and chicken (data not shown) it may indeed be a cow derived factor that could explain the farm effect.

Ecological studies are studies of risk-modifying factors on health defined either geographically or temporally. As risk-modifying factors and outcomes are averaged for the populations in each geographical unit and then compared using standard statistical methods, this study is useful for quickly generating a hypotheses as it could use already existing data. As an ecological analysis it may fail, however, for reasons already delineated in the introduction, and will need further clinical and experimental support.

At least it seems straightforward to relate IgE biology to helminthic exposure rather than to bacterial load while Fasciola may be just an indicator of mixed infections usually found in cattle. Infected cattle rarely demonstrate clinical disease, while it is known that Fasciola has numerous immunosuppressive functions in the host. IgE is not always raised (19) probably as Fasciola can degrade human immunoglobulins (20) and even induce eosinophil apoptosis (21).

So far, the possibility of any helminth effect has been excluded (22) due to the fact, that farm children do not show raised eosinophils or high IgE levels. If, however, already contact to eggs or other secreted protein is being sufficient as observed for example with Schistosoma eggs (23), this argument may not be conclusive.

This may be also the case with another recent study (24) that found remarkable differences in the asthma prevalence between Amish and Hutterite populations. The lifestyle of both communities is similar but their farming practice is distinct as the Amish follow a more traditional style of outdoor grazing whereas the Hutterities use more industrialized farming practices. Gene expression data in the Amish children have been interpreted as „intense exposure to microbes”because protection of experimental asthma by Amish derived house dust was nearly abrogated in mice deficient for MyD88. But even here the argument for any bacterial effect is not convincing as for example Schistosome egg antigen can activate both MyD88 dependent and independent pathways (24). Interestingly the gene-expression profile in the Amish children (11) look very much like the profile obtained after Fasciola infection (25).

This ecological study can not answer any of the critical questions in the introduction. It raises again the question of possible healthy worker effect as smaller herd size was a significant predictor of quitting farming (26). But even if we believe in any farm effect, cow endoparasites provide a new research strategy as DNA based identification of most helminths is now possible (27). Studies of helminth load would be needed in cattle, as well as in environmental dust, in experimental animal models using controlled exposures, as well as detailed immune profiling in humans.

Parasite effector molecules have been largely underrated so far with numerous candidates are being available for experimental and clinical testing (28). This may be even in line with the current „One Health Initiative” of the WHO addressing the connections between health and the environment by multidisciplinary research.

## Data Availability

Aggregated epidemiological data on village basis are available from the author. All other datasets are available directly online from the indicated webpages. The consent forms are no more archived.

https://geoportal.bayern.de/bayernatlas/?lang=de&topic=ba&bgLayer=atkis&catalogNodes=11,122

https://de.wikipedia.org/wiki/Oberbayern

https://www.statistikdaten.bayern.de/genesis/online/

https://edoc.ub.uni-muenchen.de/4024/1/Koch_Sandra.pdf

## Author contribution

I revised questionnaire and protocols, contacted school authorities, selected and visited all schools, examined children, supervised data entry, analyzed data, developed the hypothesis and wrote the paper.

## Acknowledgments

I wish to thank W. Lehmacher, E. Stiepel, R. Frentzel-Beyme-Bauer, B. Berbig, T. Nicolai, E. von Mutius, S. Dold, E. von Loeffelholz-Colberg and K. Winter for help with planning the study; P. Reitmeir and G. Röll for inital statistical analysis; S. Braun, A. Fuger from my team for examining the children; C. Malin, M. von Mutius, T. Kreyssig, A. Provelegios, R. Berbig, and A. Jensen for transporting our equipment; E. Kienle for technical help with the lung function devices; C. Bremer, J. Rohe, S. Funk, A. Stankiewicz, L. Pritscher, A. Wulff and M. Molette de Morangier for data typing. We thank all head teachers, school secretaries, and class teachers who participated in the project. Finally, we thank all the parents and children who took part.

During the re-analysis, many people helped with advice: P. Tschierse, C. Prazeres da Costa, T. Stöger, D. Andrade, H. Smits, J. Esser von Bieren, M. Scheuerle, L. Gershwin, G. Knubben-Schweitzer, N. Beyls, B. Gottstein and J. Charlier.

## Data availability

Aggregated epidemiological data on village basis are available from the author. All other datasets are available directly online from the indicated webpages (geoportal.bayern.de/bayernatlas, de.wikipedia.org/wiki/Oberbayern, www.statistikdaten.bayern.de/ genesis and edoc.ub.uni-muenchen.de/4024/1/Koch_Sandra.pdf)

## Conflicts of interest

I declare that I have no conflicts of interest related to this analysis. The study was conducted in 1989/1990 by order of the „Bayerische Staatsministerium für Landesentwicklung und Umweltfragen” (now „Bayerisches Staatsministerium für Wirtschaft, Landesentwicklung und Energie”) to “Dr. von Haunersches Kinderspital” (now „Kinderklinik und Kinderpoliklinik im Dr. von Haunerschen Kinderspital”).

**Supplemental Figure 1:**
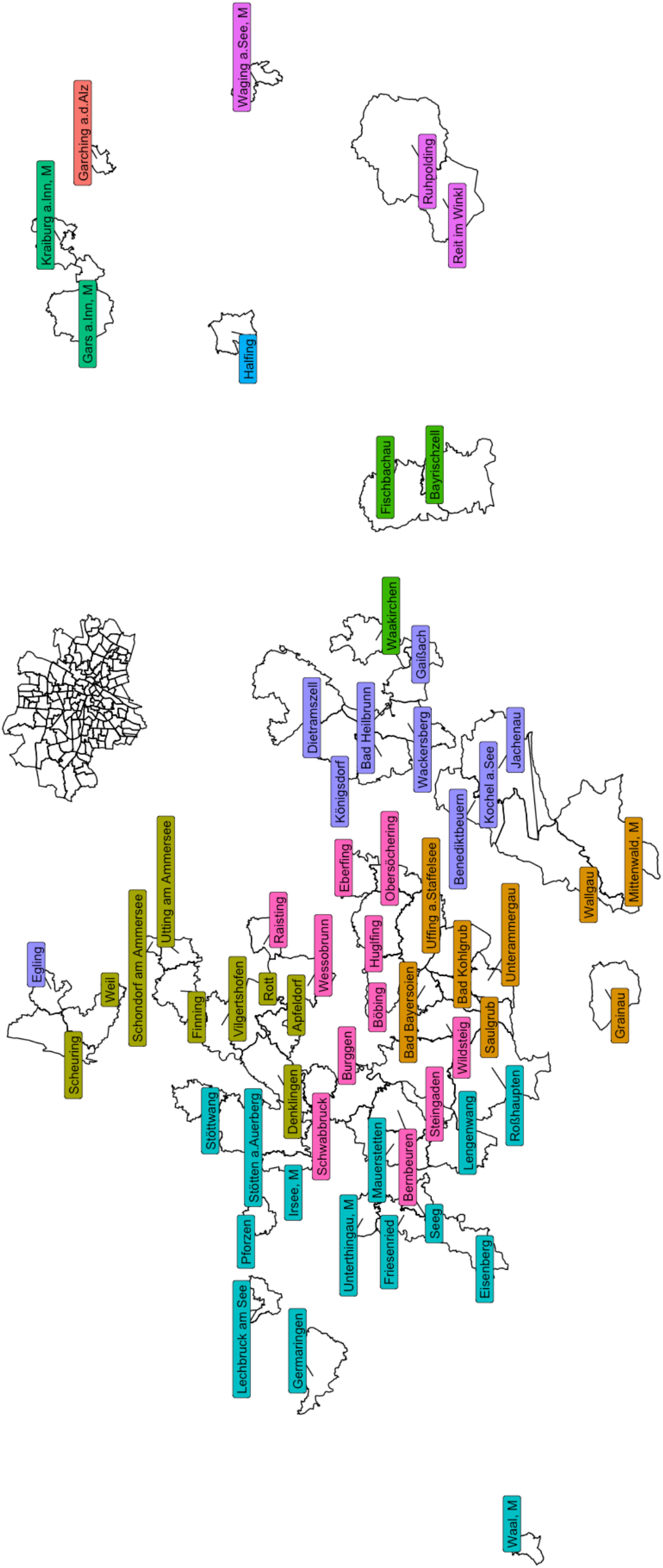
Study villages are located in the South of Munich in Upper Bavaria. For color codes see legend to Supplement Figure 2.

**Supplemental Figure 2:**
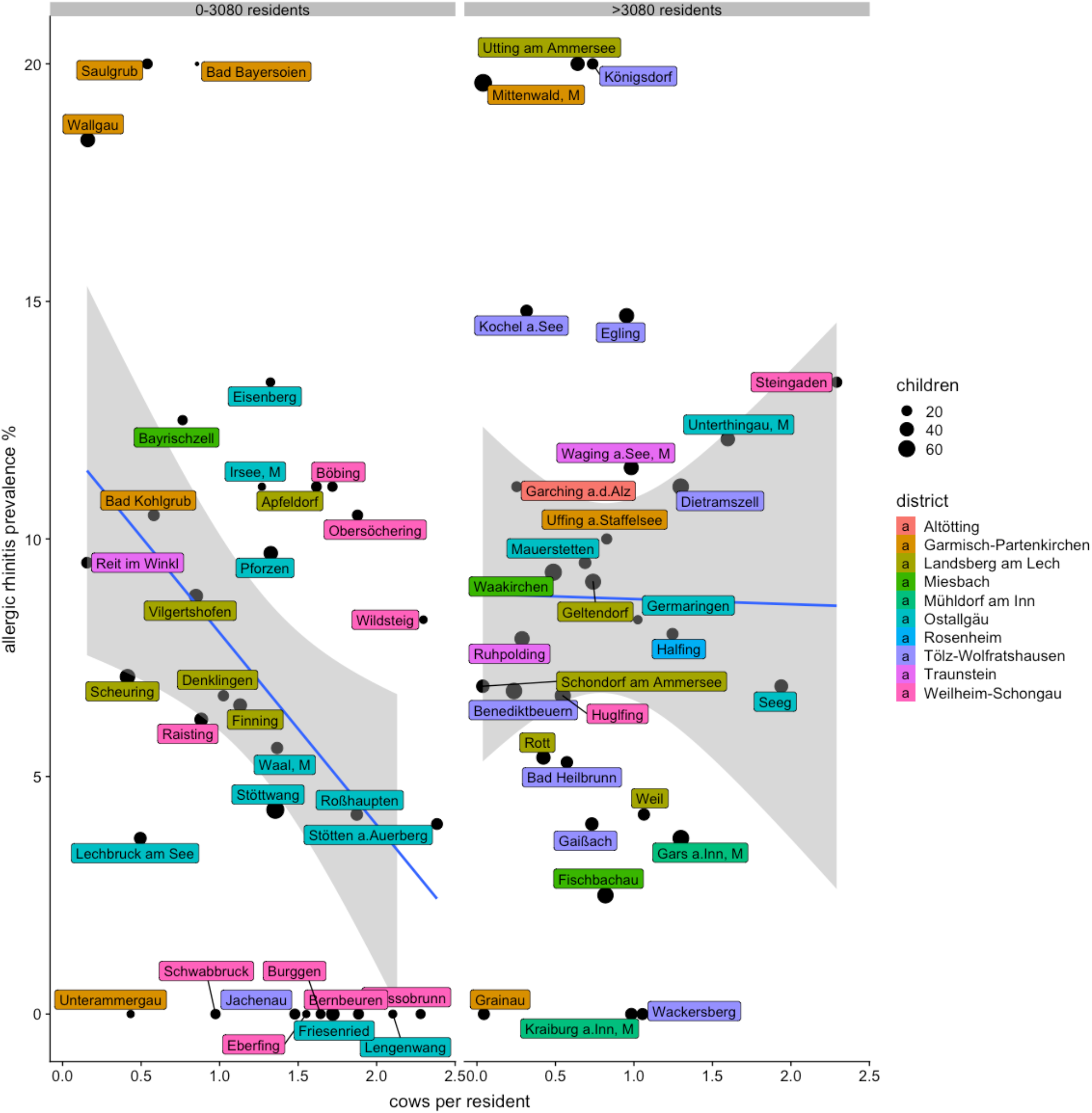
Allergic rhinitis prevalence 1989 in study villages separated by low or high resident number. Villages with allergic rhinitis >20% (outlier) are removed from the regression.

**Supplemental Figure 3:**
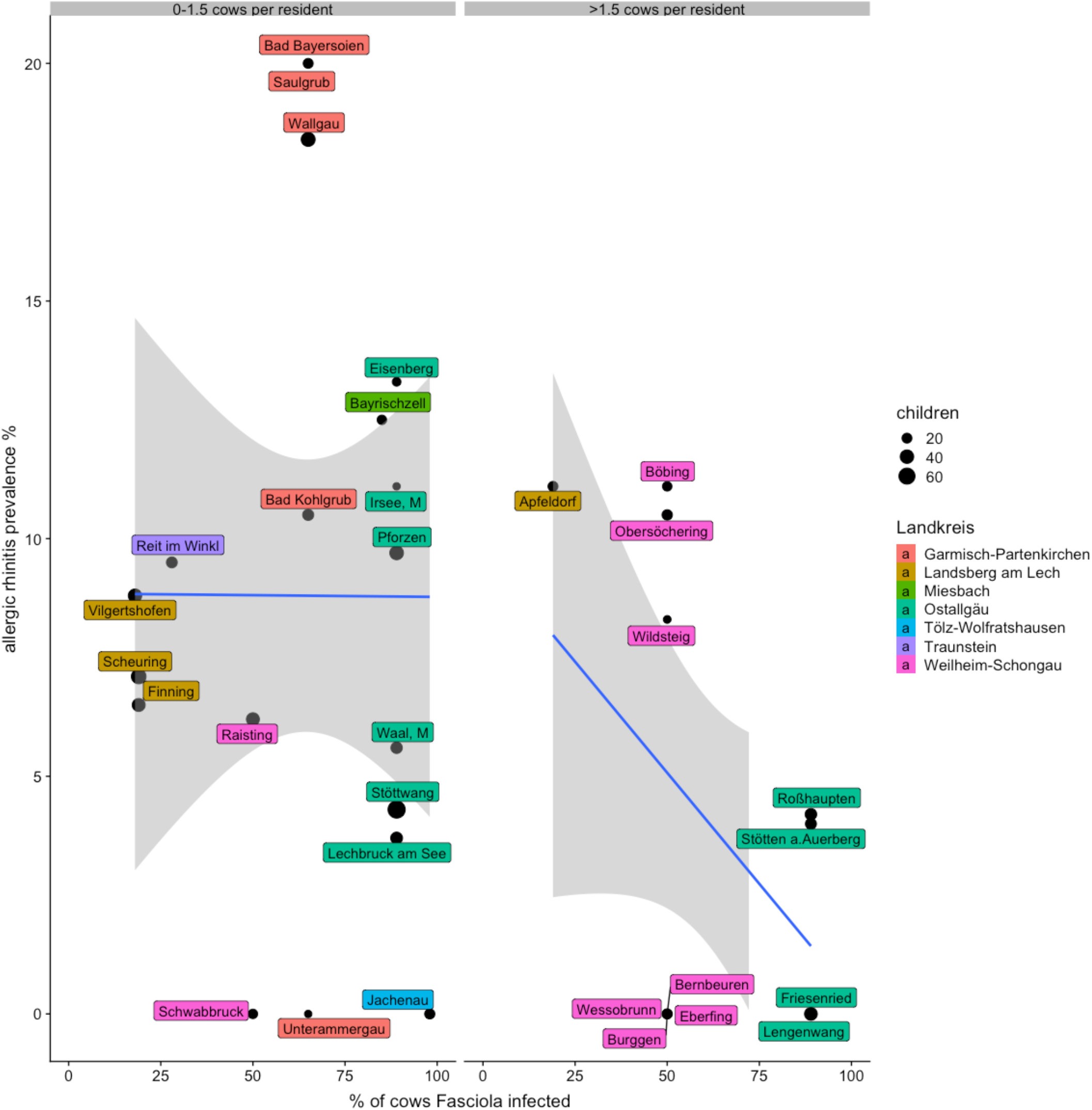
Allergic rhinitis prevalence 1989 in small sized study villages only by cattle population. Villages with allergic rhinitis >20% (outlier) are removed from the regression.

## Notes

### Competing Interest Statement

The authors have declared no competing interest.

### Author Declarations

All relevant ethical guidelines have been followed and any necessary IRB and/or ethics committee approvals have been obtained.

Any clinical trials involved have been registered with an ICMJE-approved registry such as ClinicalTrials.gov and the trial ID is included in the manuscript.

